# Using deep learning to improve genetic studies of osteoporosis

**DOI:** 10.1101/2025.09.25.25336686

**Authors:** Tore Eriksson, Chiaki Nakamori

## Abstract

To evaluate how recent advances in deep learning can improve the construction of quantitative phenotypes for genome-wide association studies (GWAS), we focused on the context of osteoporosis and bone mineral density (BMD) measurements. We applied image classifiers and transformer models to three distinct tasks. First, we developed quantitative estimates of osteoporosis severity using bone X-ray images. Second, we compared standard approaches for handling confounding variables with a multi-factor strategy based on transformer models trained on UK Biobank data. Third, we investigated whether image-based models could predict how single nucleotide polymorphisms (SNPs) associated with BMD influence bone structure. While our results were promising, application of deep learning methods did not yield substantial improvements over established approaches. Nonetheless, our findings highlight the potential of integrating imaging and machine learning techniques to refine phenotype definitions in genetic studies.

## 1 Introduction

Genome-wide association studies (GWAS) have become a cornerstone of modern genetics, leveraging the vast phenotypic and genotypic datasets generated by large-scale biobank initiatives such as the UK Biobank^1^. These resources offer extensive phenotype information across hundreds of thousands of individuals, collected through medical imaging, laboratory tests, and questionnaires. Some also provide diagnostic codes extracted from medical records, making it possible to run simple case-control GWAS analyses for well-represented diseases.

However, despite advances in electronic health records and data integration, medical datasets remain inherently incomplete due to undiagnosed conditions and untreated individuals within the population. A further limitation of using diagnostic codes in GWAS is that these binary outcomes fail to capture the severity or progression of disease. Studies have shown that GWAS using quantitative traits yield higher statistical power and improved reliability^2^.

The current standard approach for variant-to-function prediction combines epigenetic data with expression quantitative trait loci (eQTL)^3^, improving coverage but relying on tissue- or cell-specific datasets that may be unavailable or incomplete. This limitation motivates alternative strategies for interpreting genetic variants, such as direct modeling of anatomical effects using imaging data.

In this study, we apply deep learning techniques to enhance disease phenotyping, address confounding in association studies, and explore the functional impact of significant single nucleotide polymorphisms (SNPs). Our focus is on osteoporosis and bone mineral density (BMD), measured via dual-energy X-ray absorptiometry (DXA) scans. We particularly investigate whether quantitative information can be extracted from DXA images available in the UK Biobank dataset^4^.

Most of the published association studies for osteoporosis uses BMD values as a proxy for diagnoses done in the medical environment^5^. The reason for this is of course that spine BMD is a major factor in diagnosing the disease according to current clinical guidelines in most countries, including the UK^6^.

First, we examined whether imaging data could be used to derive a quantitative proxy for osteoporosis independent of clinical diagnosis. We defined initial case and control groups using clinical data, then trained EfficientNet^7^ convolutional neural networks (CNNs) to classify these groups based on single-region DXA images. For the best-performing model, we used the predicted probability of case status as a continuous GWAS phenotype, comparing the results to those from a traditional case-control GWAS.

Next, we investigated whether improved modeling of confounding variables could enhance GWAS outcomes. Traditional approaches often treat confounders such as body weight as linear covariates, despite their known non-linear effects on BMD. To address this, we constructed non-linear models incorporating sex, age, and body weight, using the residuals as adjusted phenotypes. Additionally, we trained a multitask transformer model to predict DXA-derived BMD from nearly three hundred UK Biobank phenotypes, again using residuals as phenotypes to mitigate the influence of unmeasured confounders.

Finally, we explored whether bone imaging data could be used to address the problem of elucidating the function of implied variants. In this study we removed the common step of mapping SNPs to genes, an instead tried to discern the direct anatomical effects of BMD-associated SNPs. We trained EfficientNet models to predict SNP genotypes from relevant DXA images and applied Grad-CAM^8^ to identify skeletal regions most influenced by each variant. This approach, inspired by recent work in brain imaging^9^, aims to localize the phenotypic impact of genetic variants beyond traditional statistical associations.

Although we also computed updated GWAS results for imaged UK Biobank participants, we do not delve into gene-level interpretation, as this aspect has been extensively addressed in prior studies^10–12^.

## 2 Methods

### 2.1 Individuals and phenotypes

We included participants of all ethnic backgrounds for model training, excluding only those who had formally withdrawn from the UK Biobank as of April 25, 2023.

A manual selection of physiological and environmental variables potentially influencing bone mineral density was performed using the full range of available UK Biobank data. Particular attention was given to phenotypes likely to act as confounders in BMD analyses, with a focus on categories related to diet, physical activity, and, for female participants, reproductive history.

When individuals had undergone multiple assessments with corresponding BMD measurements, each assessment was treated as a separate data point. For multiple measurements within a single assessment, values were averaged. Missing data were imputed using the mean of other assessments from the same individual when available. If no within-individual data were available, missing values were imputed using the median of participants of the same sex within a 10-year age bracket. All continuous variables were standardized to have zero mean and unit variance. We assessed the need for variable transformation by calculating the skewness of each distribution and inspecting skewed variables visually; however, no variables were ultimately log-transformed.

For DXA-derived BMD values, we used total BMD for each skeletal region, except for the femurs, where left and right measurements were treated separately. This decision was based on the assumption that measurement error is more likely than true biological asymmetry in most bones—an assumption we evaluated by examining left–right correlations (data not shown). All BMD values were centered and scaled prior to analysis.

Osteoporosis cases were identified using a combination of diagnostic codes and medication records. For ICD-9, we included code 733 (osteoporosis) and fracture codes commonly associated with osteoporotic patients of advanced age: 805 (spine), 812 (humerus), 813 and 814 (wrist), and 820 and 821 (hip). For ICD-10, we included M80 and M81 (osteoporosis), M85.8 (osteopenia), and 17 fracture-related codes from chapters S and T. Medication-based case identification included prescriptions for raloxifene hydrochloride (a selective estrogen receptor modulator), teriparatide (a parathyroid hormone analog), denosumab (a RANKL inhibitor), calcitonin, and ten bisphosphonate formulations. Participants with either a qualifying diagnosis or medication were classified as osteoporosis cases; all others were considered controls.

### 2.2 Image-based osteoporosis prediction

As described above, participants with either a qualifying diagnosis or osteoporosis-related medication were labeled as cases, while all others were considered controls. For each anatomical image region, we trained a binary classifier using the EfficientNet-V2S architecture to distinguish between cases and controls.

All images were preprocessed by inverting grayscale values and cropping to a uniform size of 672×672 pixels, except for whole-body images, which were initially cropped to 300×900 pixels and then resized to 672×672. Each image was processed using a single grayscale channel.

The dataset was split into training, validation, and test sets, with 10% of the images reserved for testing and 9% for validation. Models were trained using early stopping with a maximum of 50 epochs and a batch size of 12. Optimization was performed using the Adam optimizer with a learning rate of 0.0005, while all other hyperparameters were kept at their default values.

### 2.3 Non-linear model of confounding variables

Non-linear models were evaluated using the `lmer` function from the **lme4**package^13^in R. Several models incorporating age, weight, and sex were compared. To improve interpretability, age and weight were centered at baseline values of 40 years (*Age40*) and 75 kg (*Weight75*), respectively.

Based on visual inspection of residual plots, we selected a model in which *Age40*was modeled as a linear term stratified by sex, and *Weight75*was modeled as a second-degree polynomial. The final model specification is shown in Eq. 1:

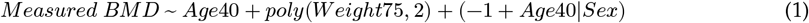

### 2.4 Transformer model

A multi-task encoder model was implemented in Python, based on the original transformer architecture described by Vaswani *et al*.^14^. Numerical phenotypes were projected into the model’s hidden dimension using dedicated dense layers, while categorical variables were embedded using variable-specific `Embedding` layers from the **Keras**library^15^. The encoder outputs were aggregated via average pooling and passed through a fully connected layer with leaky ReLU activation, followed by a final dense layer with linear activation to produce 25 output values.

Hyperparameters were tuned by splitting the dataset evenly into training and evaluation sets. The final model used a hidden dimension of 128 and consisted of four transformer layers, each with eight attention heads. Training was performed using the Adam optimizer for up to 100 epochs, with early stopping and a scheduled learning rate.

After tuning, the final model was retrained on the full dataset and used to predict BMD values. Individual-level residuals were computed for all participants and used as adjusted phenotypes in downstream analyses.

### 2.5 Genome-wide analysis

Sample quality control (QC) and selection of British participants were performed as previously described^16^. Using QC data provided by the UK Biobank, we excluded individuals meeting any of the following criteria: (1) heterozygosity outliers or missing heterozygosity data, (2) kinship inference status marked as “exclusion”, (3) more than ten third-degree relatives, (4) not included in the principal component analysis (PCA) calculation, or (5) non-standard karyotypes (*i.e*., not XX or XY).

To define a genetically homogeneous British cohort, we computed a deviation score D_i_for each individual based on the first six genetic principal components (PCs), using the formula in Eq. 2. Here, 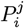 denotes the PC score for individual *i*and component *j*, while M^j^and SD^j^represent the mean and standard deviation of each PC among individuals self-identifying as “British” or “Irish”. Individuals with D_i_< 7^2^were retained for downstream analysis.

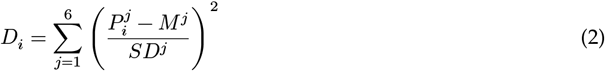

PCA was recomputed using the selected British samples to generate updated PC scores for association testing. SNPs located in long-range linkage disequilibrium (LD) regions were excluded^17^. PCA was performed using PLINK^18^on SNPs with minor allele frequency (MAF) > 0.05 and pairwise LD r^2^< 0.05.

Genome-wide association analyses were conducted using the mixed-model method implemented in BOLT-LMM (v2.4)^19^. SNPs with imputation quality scores > 0.9 were filtered based on MAF > 0.001 and Hardy–Weinberg equilibrium p-values > 1×10^-5^in the final population using PLINK. To select representative SNPs for model fitting, we applied LD pruning using a 500 kb window and an r^2^threshold of 0.1 (--indep-pairwise 500 10 0.1). These pruned SNPs were used for model training, while the remaining SNPs were included as imputed variants in the association tests.

For residual-based phenotypes, genotyping array and the first four genetic PCs were included as covariates. For measured BMD values, sex, age, and weight were also included. LD scores from the 1000 Genomes Project European (EUR) population were used for LD score regression.

### 2.6 SNP selection

Due to the computational cost of training models on all genome-wide significant SNPs, we selected a representative subset covering approximately ten association peaks per DXA region. SNPs were first prefiltered using a significance threshold of P < 1 × 10^-5^. These SNPs were then grouped into genomic blocks of one million base pairs, and the 12 blocks with the lowest minimum P-values were selected for each region.

Because DXA measurements do not map one-to-one onto specific imaging regions, we manually assigned each DXA region with corresponding anatomical image regions, as summarized in Table 1. SNPs associated with the L1–L4 spine region were also tested using Lateral Vertebral Assessment (LVA) images.

**Table 1:** Correspondence of DXA measurements with DXA image regions.

| DXA Phenotype | Region |
| --- | --- |
| Head | Total_Body |
| Arms | Total_Body |
| Ribs | AP_Spine |
| Ribs | Total_Body |
| Trunk | Total_Body |
| L1-L4 | LVA |
| Spine | AP_Spine |
| Pelvis | Left_Femur |
| Pelvis | Right_Femur |
| Pelvis | Total_Body |
| Femur_lower_neck-left | Left_Femur |
| Femur_lower_neck-right | Right_Femur |
| Femur_neck-left | Left_Femur |
| Femur_neck-right | Right_Femur |
| Femur_shaft-left | Left_Femur |
| Femur_shaft-right | Right_Femur |
| Femur_total-left | Left_Femur |
| Femur_total-right | Right_Femur |

Table 1: Correspondence of DXA measurements with DXA image regions
| DXA Phenotype | Region |
| --- | --- |
| Femur_upper_neck-left | Left_Femur |
| Femur_upper_neck-right | Right_Femur |
| Femur_upper_neck-left | Left_Femur |
| Femur_upper_neck-right | Right_Femur |
| Femur_wards-left | Left_Femur |
| Femur_wards-right | Right_Femur |
| Legs | Left_Ortho_Knee |
| Legs | Right_Ortho_Knee |
| Legs | Total_Body |
| Total | Total_Body |

To further reduce redundancy caused by overlapping association peaks across GWAS runs, we selected a single representative SNP per region using the `--indep-pairwise` command in PLINK 2.0. This was performed with a window size of 10 Mb and an LD threshold of r^2^< 0.9, ensuring that selected SNPs were not in high linkage disequilibrium with one another.

### 2.7 Image classification by genotype

The genotype classification models followed a similar architecture to the osteoporosis prediction models, but used the more lightweight EfficientNet-B0 variant to reduce training time. Consequently, input images were resized to 224×224 pixels. To improve model performance, we first pretrained a base model on the task of predicting the sex of the depicted individual and then used the learned weights to initialize the genotype classification model.

We compared two approaches for genotype prediction: categorical classification of the three genotype classes and ordinal regression based on allelic dosage. Based on training progression and performance (data not shown), we selected the categorical classification approach.

Saliency maps were generated for each model using TensorFlow’s gradient tape functionality. Gradients from multiple convolutional layers were averaged to produce heatmaps highlighting image regions most influential for genotype prediction.

3 Results

### 3.1 Selection of individuals

A total of 429,015 participants of genetically confirmed British or Irish ancestry were selected, using selfreported ethnicity as a reference. Among these, 26,790 individuals (13,401 females and 13,389 males) had available DXA imaging data. The distribution of age and body weight within this subset is illustrated in Fig. 1.

**Figure 1.**
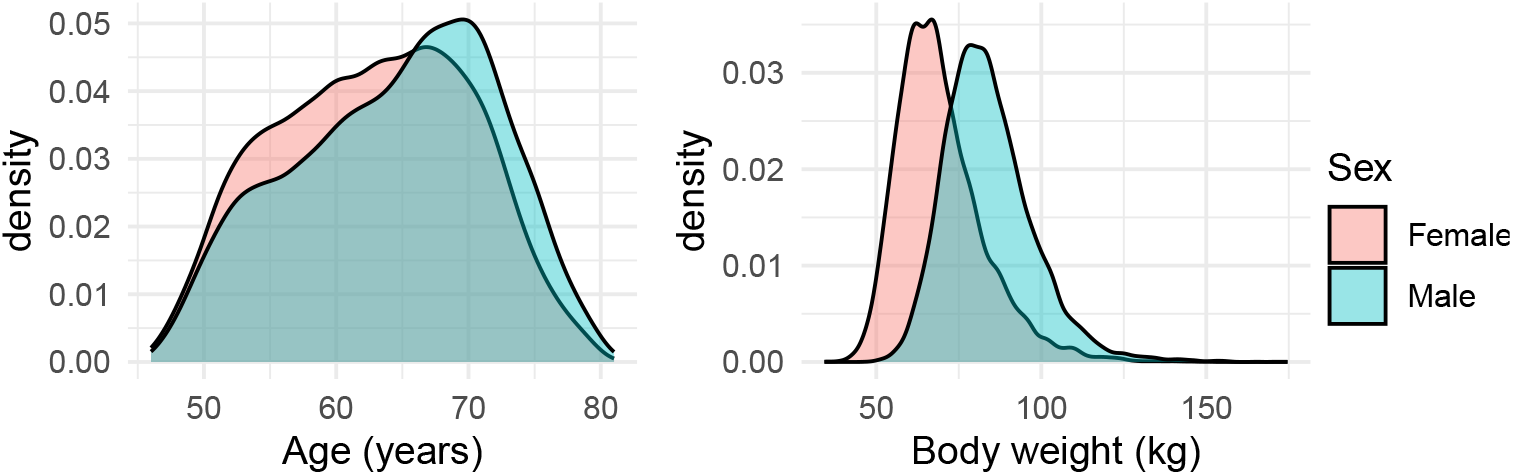
Age and body weight distributions by sex.

### 3.2 Osteoporosis prediction

The final dataset used for osteoporosis prediction included 1,342 cases and 9,142 controls. We assessed the predictive performance of each region-specific model by generating receiver operating characteristic (ROC) curves using the **pROC**package^20^in R (Fig. 2), excluding images used during model training.

**Figure 2.**
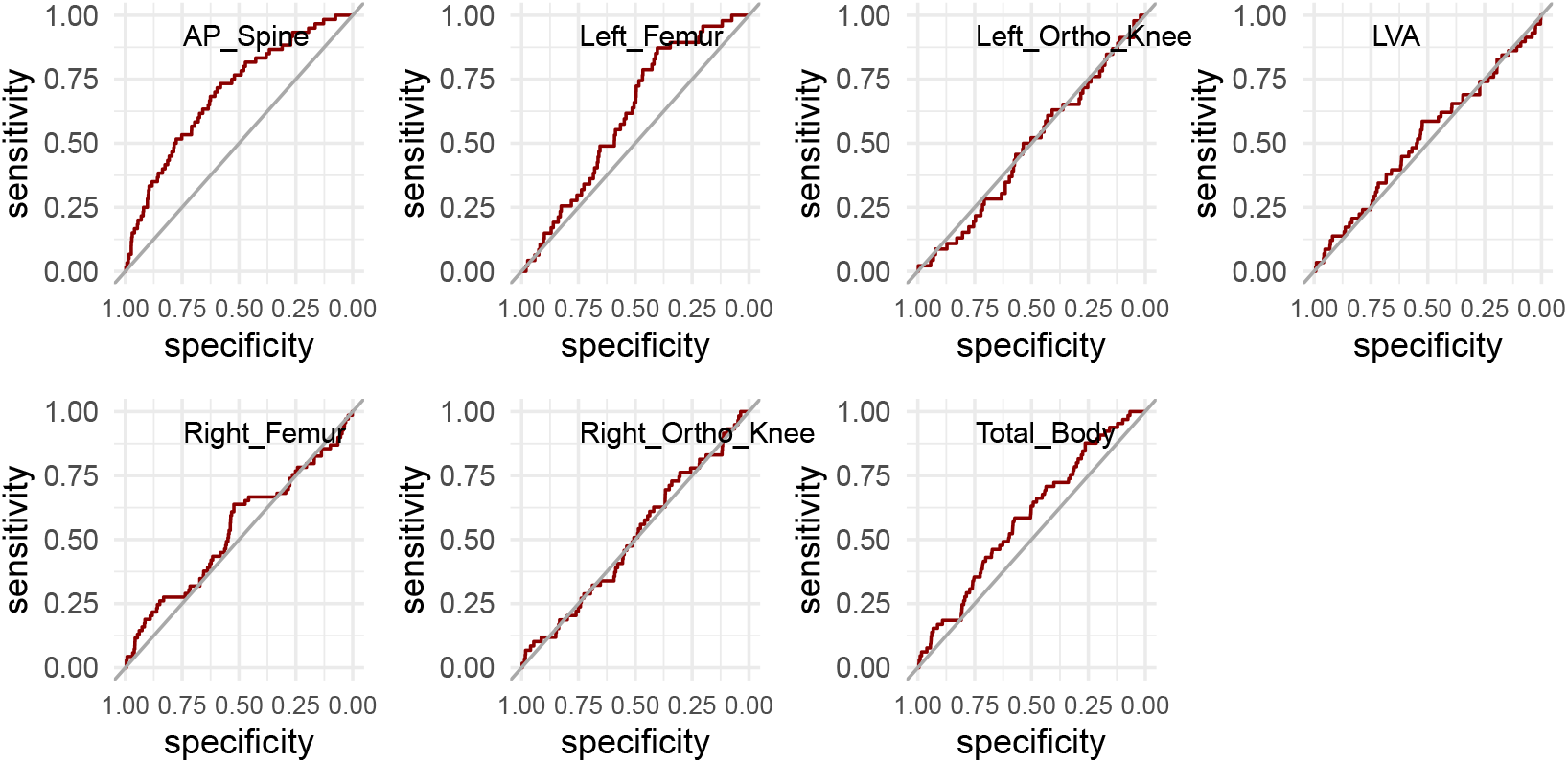
ROC curves for each model. The straight line represents a model without predictive capability.

The model based on anterior-posterior spine (AP_Spine) images demonstrated the highest performance, with an area under the curve (AUC) of 0.698. In contrast, most other models failed to effectively distinguish cases from controls using DXA images. Predicted values for cases and controls from the AP_Spine-based model are presented in Fig. 3.

**Figure 3.**
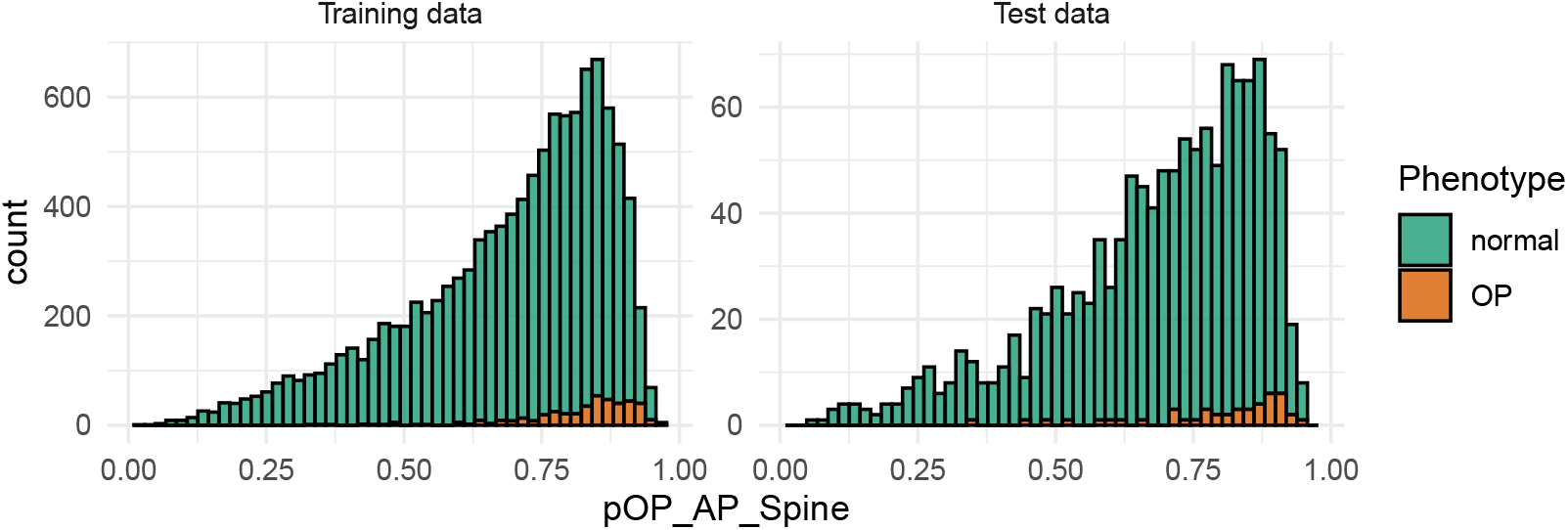
Predicted probabilities for osteoporosis for the model trained on anterior-posterior spine DXA images. Images used in training are displayed on the left, others on the right.

A comparison between the GWAS results based on osteoporosis case-control status (Fig. 4a) and those derived from the AP_Spine-based prediction model (Fig. 4b) reveals notable differences in the distribution of genetic signals. The AP_Spine-based model produced clearer and more distinct association peaks, suggesting potentially stronger or more localized genetic contributions. However, this observation was not formally quantified, and further statistical evaluation would be necessary to confirm the extent and significance of these differences.

**Figure 4.**
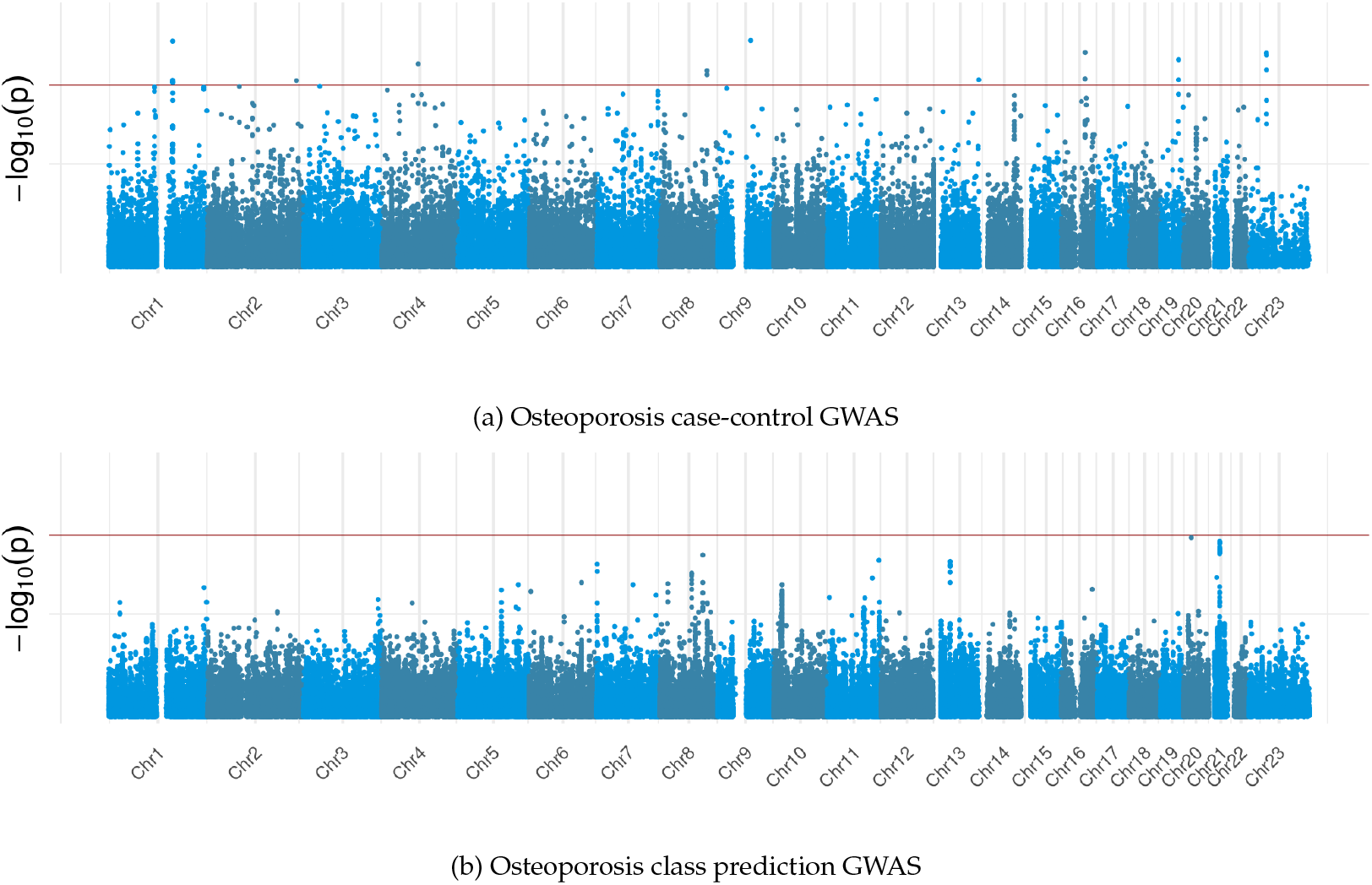
GWAS Manhattan plots for diagnosed and predicted osteoporosis.

### 3.3 Confounding variables

Representative effects of two approaches to handling confounding variables are illustrated using data from the pelvis and legs. In the pelvis region (Fig. 5), we observe that the non-linear model does not fully eliminate the influence of body weight in male participants. This residual confounding suggests that the model may require further refinement or additional covariate adjustment to more effectively isolate bone-specific signals.

**Figure 5.**
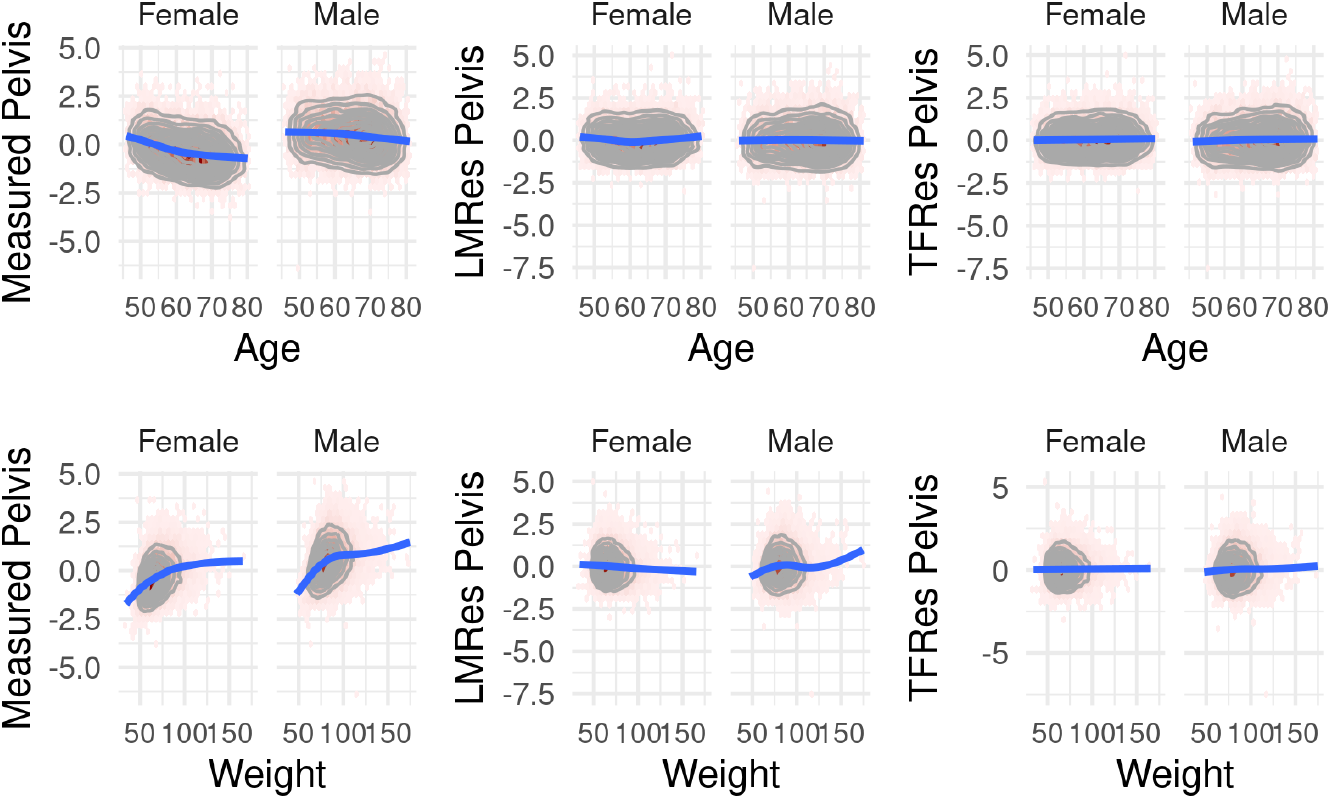
Measured DXA and model residuals for pelvis.

A similar residual effect of body weight is observed in the legs (Fig. 6), along with additional sex-specific differences in the influence of age on leg bone mineral density.

**Figure 6.**
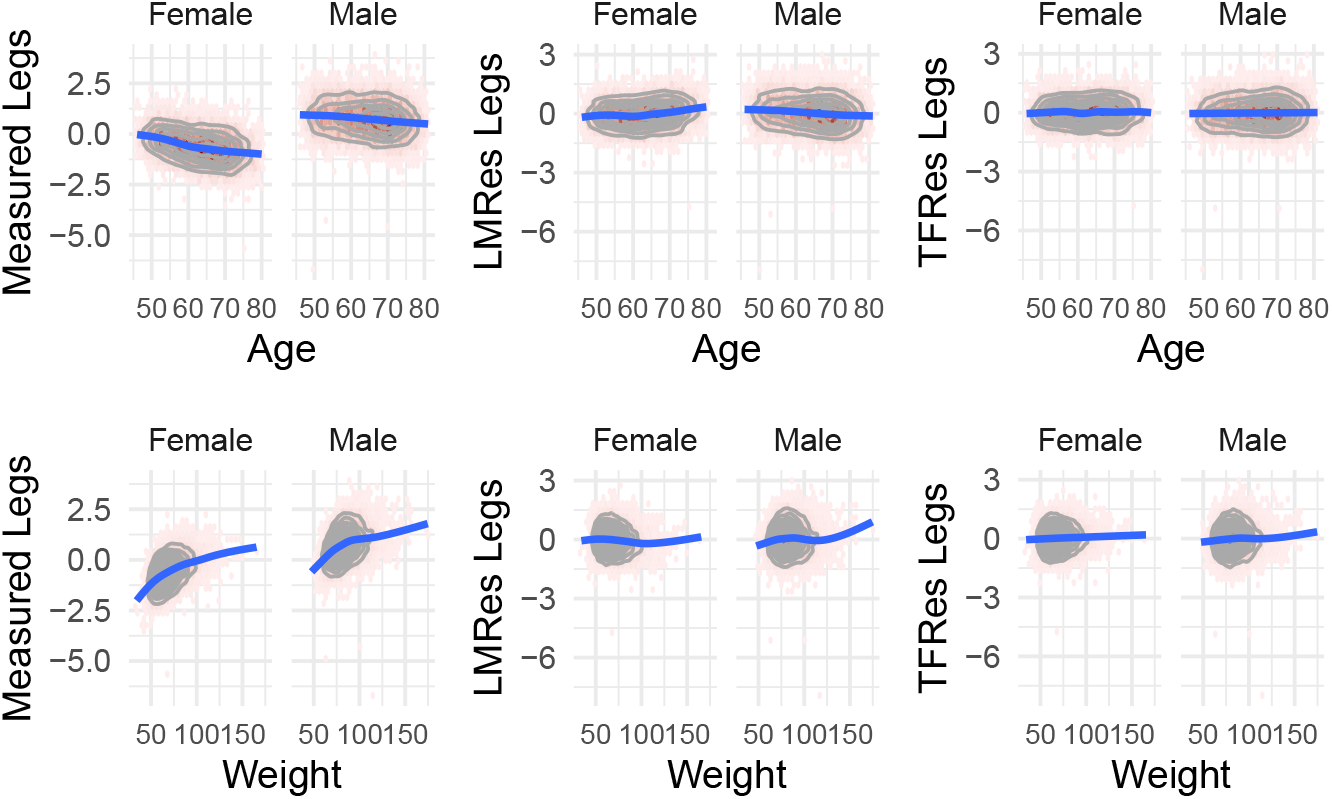
Measured DXA and model residuals for legs.

To further evaluate the impact of confounding adjustment methods, we compared the GWAS results across the three modeling approaches. A representative example is shown in the Manhattan plots for the leg region (Fig. 7). Overall, differences between the methods were modest, with p-value variations typically within one order of magnitude, indicating that the choice of confounding adjustment method had limited impact on the detection of genetic associations in this context.

**Figure 7.**
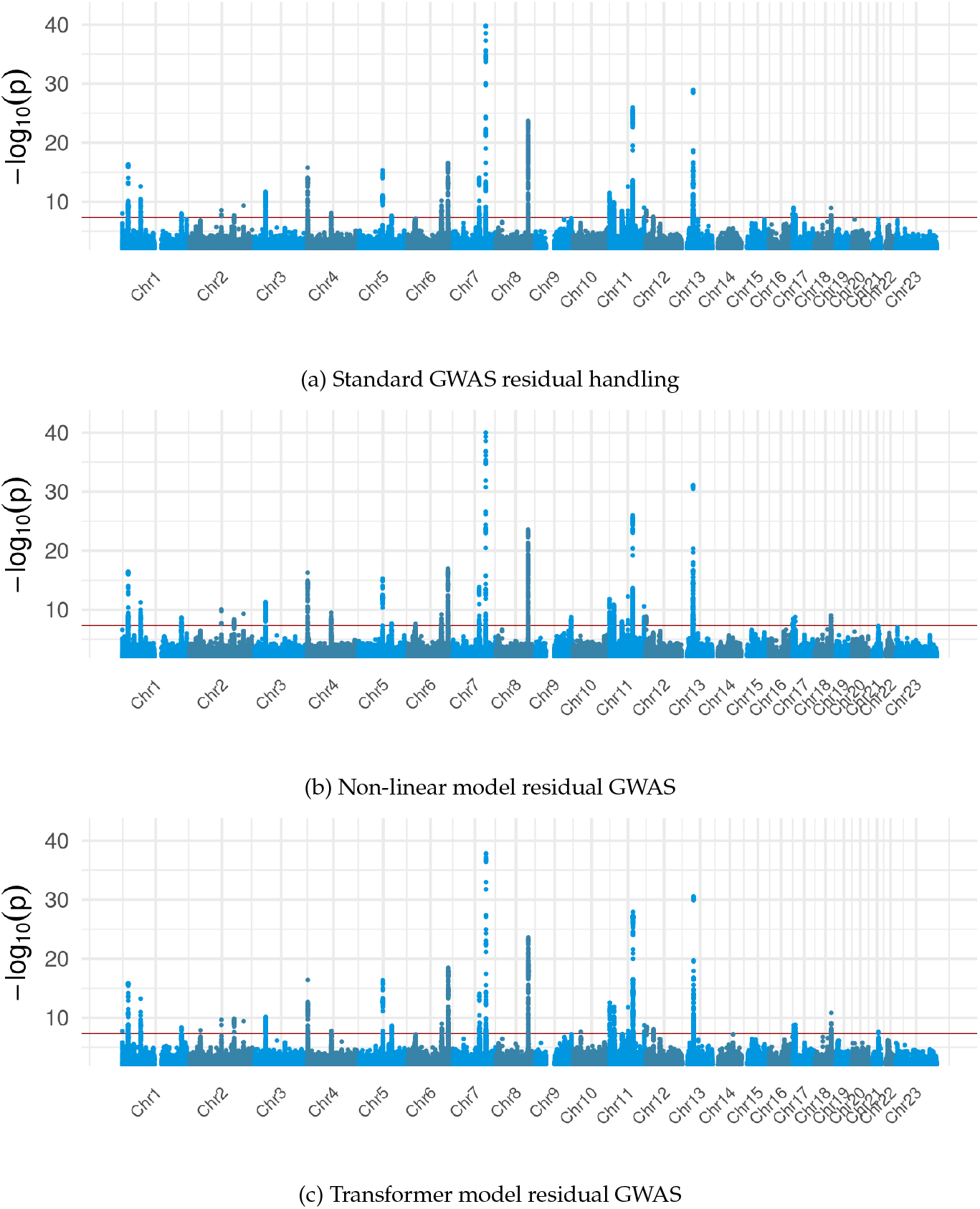
Comparison of GWAS results based on leg DXA BMD. Methods for dealing with confounding variables are standard, non-linear model, and transformer model, from top to bottom. Genome-wide significance is described by a red line at a nominal p-value of 5×10^-8^.

### 3.4 SNP selection

After identifying approximately ten GWAS peaks from each anatomical region and subsequently performing linkage disequilibrium clumping, we obtained a total of 186 region-SNP combinations. The femur region contributed the largest number of SNPs, likely due to the availability of DXA measurements across multiple subregions (Fig. 8).

**Figure 8.**
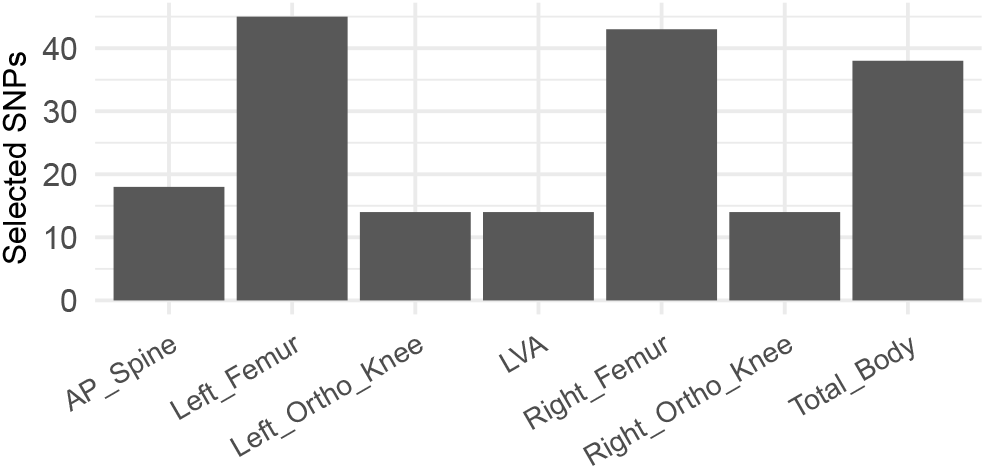
Number of selected SNPs by anatomic region.

The SNP most frequently selected across regions was rs3779381 which appeared in six anatomical regions. This variant is located in the first intron of the *WNT16*gene, a well-established regulator of bone mineral density. Notably, rs3779381 showed consistent correlations with BMD across all examined regions (Fig. 9), reinforcing its potential as a key genetic determinant of skeletal health.

**Figure 9.**
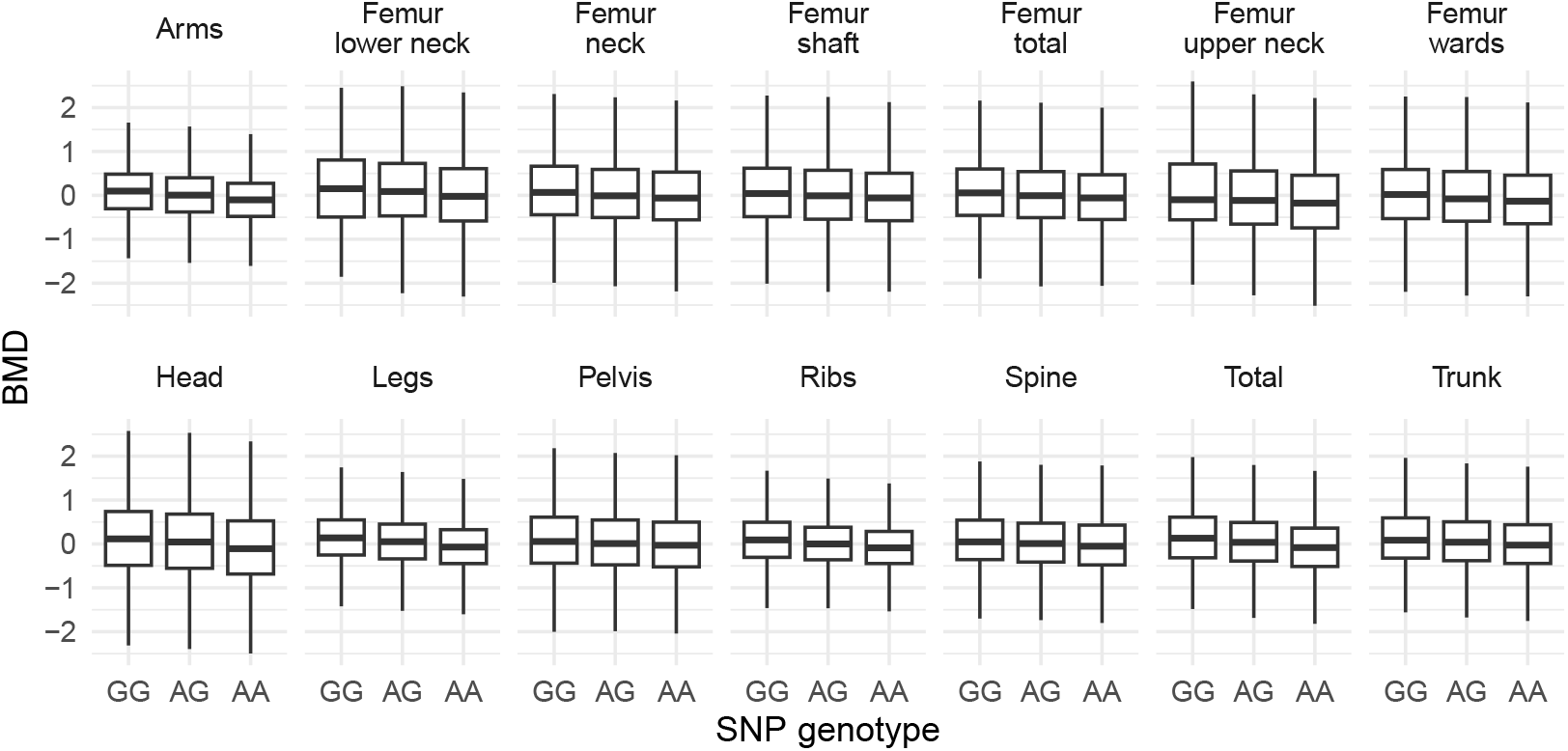
rs3779381 genotype correlation with transformer-corrected bone mineral densities.

### 3.5 Genotype prediction

Attempts to train models for genotype prediction using DXA images were unsuccessful. However, the pretrained models showed strong performance on the task of predicting sex, with all models achieving an area under the curve of 0.98 or higher on non-training data. The lowest AUC was observed for the Lateral Vertebrae Assessment images, at 0.984.

Grad-CAM visualizations of the sex-predictive models revealed consistent anatomical regions contributing to classification. For whole-body images (Fig. 10a, Fig. 10b), the models predominantly used the rib cage and pelvis to identify females, and the elbow and knee regions to identify males. In femur-specific models, female classification appeared to rely on the shape of the ilium, particularly near the ischial spine, while male classification focused on the femoral neck and head (Fig. 10c, Fig. 10d).

**Figure 10.**
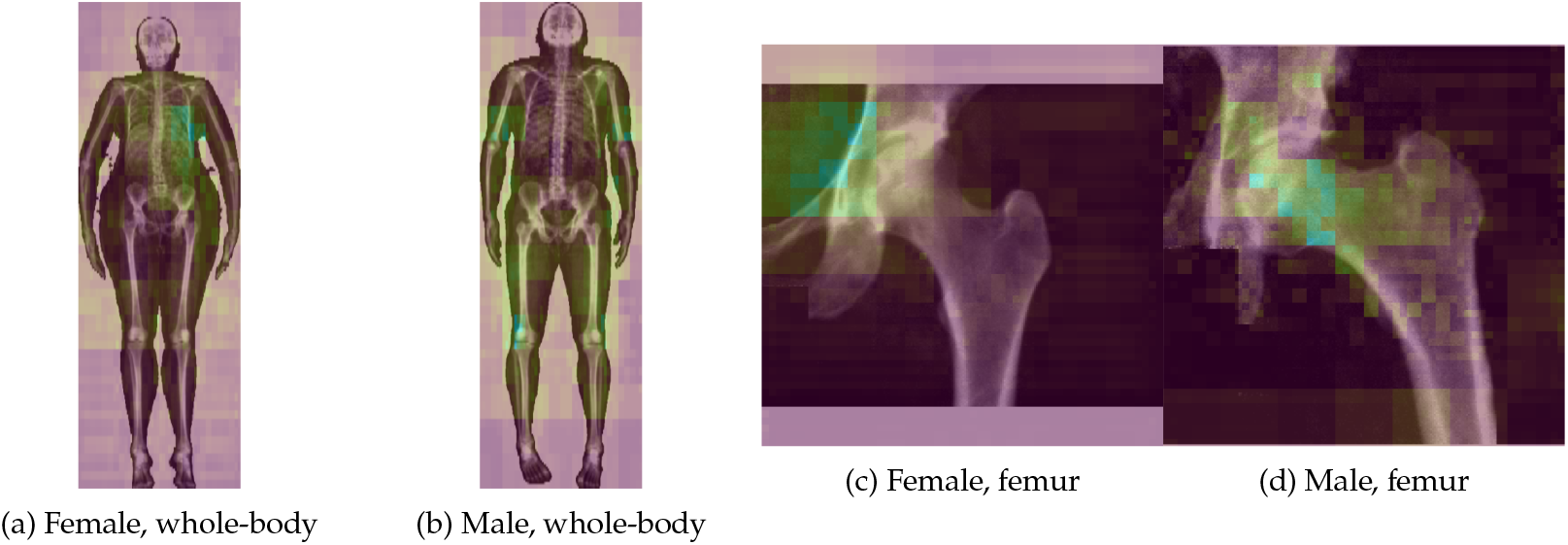
Example Grad-CAM heatmaps when predicting sex from whole-body or femur images.

While some genotypes initially showed promising validation accuracy during training, further inspection revealed that these variants shared a low minor allele frequency, typically below 1%. This suggests that the apparent learning may have been driven by overfitting to rare patterns rather than meaningful generalization.

To investigate whether image resolution was a limiting factor, we tested a larger model architecture (V2S) using higher-resolution DXA images (672 × 672 pixels). This model was applied to the SNP rs3755955, which had exhibited the most encouraging learning curve in earlier experiments. However, training the model for 100 epochs did not yield any improvement in predictive performance (data not shown), indicating that increased image resolution alone may not be sufficient to overcome the challenges of genotype prediction from DXA images.

## 4 Discussion

One of the primary objectives of this study was to ascertain whether imaging data could provide a quantitative proxy for osteoporosis, independent of clinical diagnoses. Our results demonstrate that an EfficientNet CNN trained on anterior-posterior spine (AP_Spine) DXA images effectively distinguished osteoporosis cases from controls, achieving an AUC of 0.698. While most other anatomical regions yielded less effective models, the performance of the AP_Spine model is encouraging. Notably, GWAS conducted using the continuous phenotype derived from this AP_Spine-based prediction model revealed clearer and more distinct association peaks compared to traditional case-control GWAS. This suggests that deep learning-derived phenotypes may enhance the detection and localization of genetic signals, potentially leading to a deeper understanding of genetic contributions to osteoporosis. However, the extent and significance of these differences require formal quantification and further statistical evaluation.

We also investigated methods to use deep learning to improve the modeling of confounding variables, such as age, sex, and body weight, which are known to have non-linear effects on BMD. Using non-linear statistical models and a multi-task transformer model to predict DXA-derived BMD, we were able to remove a lot of the effect of confounding. Despite this, a comparison of GWAS results across standard, non-linear, and transformer models showed only modest differences, with p-value variations typically within one order of magnitude. This indicates that, in the context of detecting genetic associations for BMD, the choice of confounding adjustment method had a limited overall impact.

In previous projects involving PheWAS analyses with biobank data, we have also encountered the critical need for robust methods to map genome-wide association peaks to their corresponding effector genes—a process known as variant-to-function prediction. Early approaches focused primarily on variants located in promoter or transcribed regions, but these missed many regulatory relationships, especially for peaks overlapping enhancers located far from the target gene.

Our current standard approach combines epigenetic data with expression quantitative trait loci using colocalization analysis^21^overlaid on epigenetically active regions in relevant cell types. This approach has significantly improved the explainability of detected GWAS peaks. However, this method depends on the availability of epigenetic data from tissues and cell types relevant to the phenotype, ideally from a similar population. In many cases, such data may be limited or unavailable.

Thus, a novel aspect of this study was the attempt to discern the direct anatomical effects of BMD-associated SNPs from DXA images, bypassing the step of mapping SNPs to genes. The aim was to localize the phenotypic impact of genetic variants in the images using Grad-CAM. We trained EfficientNet image models to predict a number of SNP genotypes from relevant DXA images covering all regions available. Despite these efforts, our attempts to train models for genotype prediction directly from images were unsuccessful. Apparent learning for some variants was often driven by overfitting to rare patterns, particularly for SNPs with low minor allele frequency (MAF < 1%). Utilizing larger model architectures also did not yield improvements in predictive performance. This suggests that directly predicting specific genotypes from raw DXA images may be inherently challenging or require different imaging modalities or deep learning architectures not explored here.

## 5 Limitations

Medical datasets, including those from large biobanks, are inherently incomplete due to undiagnosed conditions and untreated individuals. Our initial attempts to detect undiagnosed vertebral compression fractures or derive additional bone phenotypes directly from DXA images were unsuccessful, underscoring the complexity and current limitations of extracting detailed diagnostic information from these images.

While our image-based models successfully predicted straightforward phenotypes such as sex, and also captured a signal for osteoporosis, this result is expected given that low lumbar spine BMD is a key diagnostic criterion for osteoporosis^6^. However, we were unable to extract a strong genetic signal directly from the images. This limitation may reflect the modest influence of individual variants on bone phenotype. Other contributing factors could include the relatively low resolution of the DXA images, the use of compact image models, and the unbalanced training data for variants with low minor allele frequency.

Future work may benefit from access to higher-resolution imaging data, more expressive model architectures, and improved strategies for handling rare variants. These enhancements could enable more accurate phenotype extraction and variant interpretation from medical images.

## 6 Conclusion

The present study leverages deep learning techniques and extensive UK Biobank data to advance osteoporosis phenotyping, refine association studies, and explore the functional impact of single nucleotide polymorphisms on bone mineral density. Our findings offer valuable insights into the utility of imaging data for identifying disease-related traits and genetic associations, while also highlighting key challenges in extracting certain types of genetic information directly from images.

## Data Availability

The data and code used in this study are available through the UK Biobank Access Management System (AMS) under Application Number 74307. Access to the data is subject to approval by UK Biobank.

## 7 Author contributions

Conceptualization, T.E.; Data Curation, T.E., C.N.; Investigation, T.E.; Methodology, T.E. and C.N.; Software, T.E. and C.N.; Visualization, T.E.; Writing, T.E. and C.N.; Resources, T.E.; Supervision., T.E.

## 8 Acknowledgment

This research has been conducted using the UK Biobank^1^Resource under Application Number 74307. Microsoft Copilot was used to improve text flow and grammar.

